# Correlation between PCR Examination Rate among the Population and the Containment of Pandemic of COVID-19

**DOI:** 10.1101/2020.05.13.20100982

**Authors:** Masahiro Sonoo, Masashi Idogawa, Takamichi Kanbayashi, Takayoshi Shimohata, Hideyuki Hayashi

## Abstract

We investigated the relation between PCR examination rate among population and the success of containment of COVID-19 for each country. Although there was moderate correlation, multiple regression revealed that the success of containment was solely explained by Gross Domestic Product per capita (GDP), which may well be related to the strict compliance to social distancing. Close inspection of individual countries supported this hypothesis. The social distancing must be the largest factor to achieve containment, and the contribution of broad PCR tests is small.

---

Coronavirus disease 2019 (COVID-19) pandemic is now a worldwide peril and its control is an emergent issue. Expansion of PCR tests is generally believed to be the most important key for containing the infection,^1^ together with social distancing. However, the relation between examination rate and the success of containment has not been directly studied to our knowledge. We investigated this issue based on the open data at websites.^2,3^

The rate of PCR tests among population (named as Examination Rate; ER) was extracted from a website for countries having more than 1000 cases.^2^ To evaluate the success of containment, the latest value of the new cases per 1 million population during 7 days in the trajectory analysis^3,4^ was divided by its highest value, and was named as the Containment Ratio (CR). We postulated that strict compliance to social distancing is better achieved in advanced countries, and therefore Gross Domestic Product per capita (hereafter GDP) of each country was adopted as another predictor variable. Countries lacking some data were excluded. For instance, China was not included because ER is not known. The correlation between CR and ER/GDP was investigated by simple and multiple regressions/correlations. All statistical calculation was done using Microsoft Excel for Macintosh.

Included were 90 countries. Individual countries are plotted in Figure 1 using logarithmic scales for every parameter. Notable countries are marked. As results, CR was negatively correlated both with ER (r = –0.423, p < 0.0001) and GDP (r = –0.483, p < 0.00001). In multiple regression, p-values of partial regression coefficients were 0.336 for ER and 0.009 for GDP, only the latter was significant. In other word, although CR was negatively correlated with ER, this was thought to be due to confounding effect with GDP (ER and GDP were highly correlated, r = 0.752). Accordingly, it is concluded that CR is explained by GDP alone.

**Figure 1.**
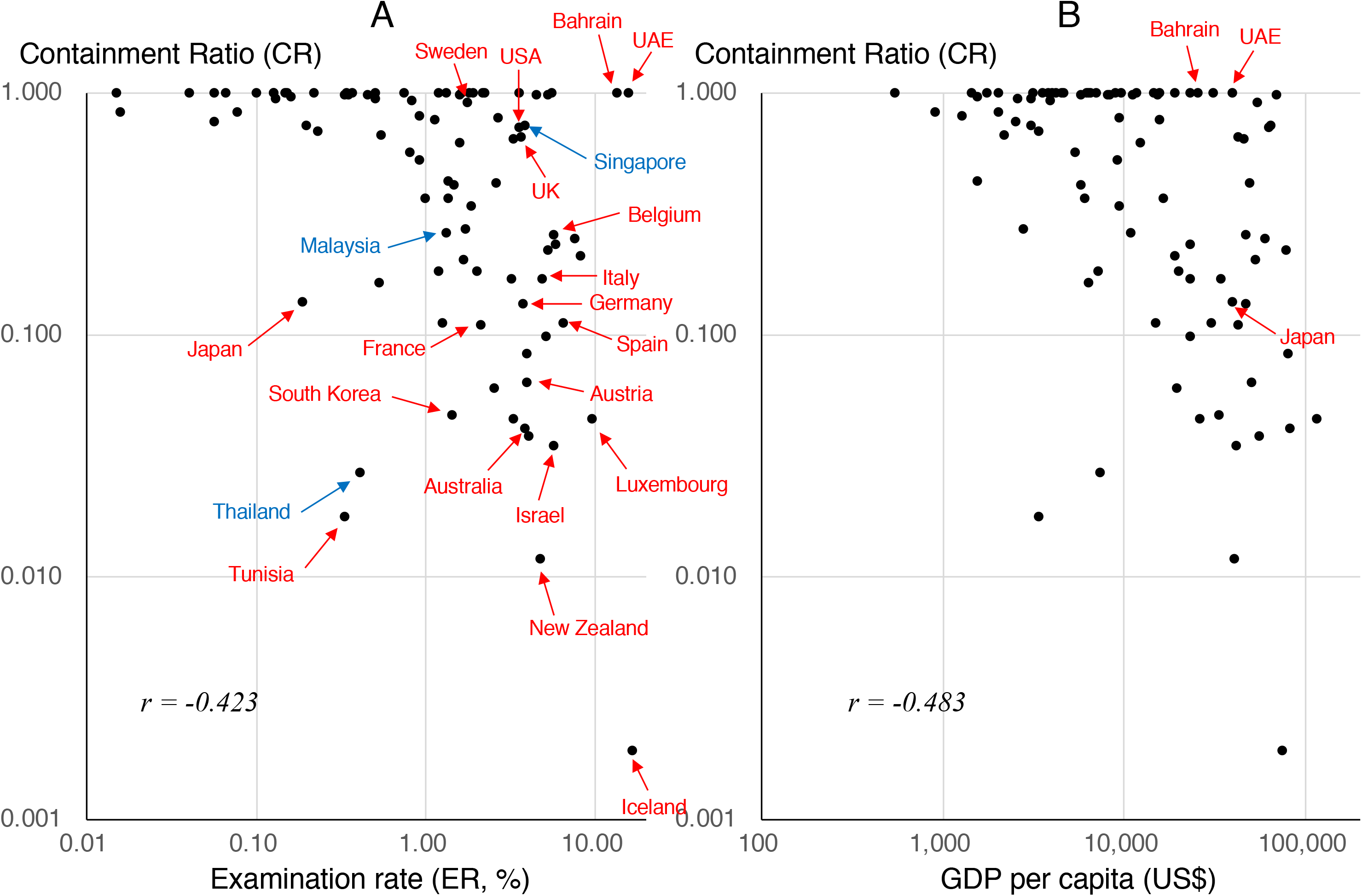
Relation between Containment Ratio (CR) vs. Examination rate (ER) or GDP. A: CR vs. ER B: CR vs. GDP Notable countries are marked. GDP was acquired from https://www.globalnote.jp/p-datag/?dno=8870&post_no=1339 Accessed May 1, 2020. Abbreviations: ER, Examination Rate; CR, Containment Ratio; GDP, Gross Domestic Product per capita.

Close inspection of individual countries reveals several interesting facts. Countries with the three highest ER in the world are Iceland, UAE, and Bahrain. Iceland achieved world best containment, but in UAE and Bahrain the pandemic is not at all controlled. This may be related to large population of foreign workers in these countries. Three South-East Asian countries, Thailand, Malaysia and Singapore having similar climate and economical states (blue marks in Figure) showed a “positive” correlation, i.e. opposite to the general tendency. Singapore achieved very high ER (3.8%) among Asian countries but CR remains high. This may be related to the fact that Singapore government discouraged healthy peoples to wear a mask until early April.^5^ However, this strategy may not have been so bad since attenuation of virulence is reported in Singapore,^6^ and our analysis predicted the lowest Infection Fatal Rate (0.005%) in the world in Singapore (unpublished results). The good result of Thailand may be due to prompt and intensive preventive measures including lock-down.^7^ Japan is criticized by its very low ER (0.19%).^8^ However, when compared with major western countries in north hemisphere generally achieving 2 to 7% ER, its CR is almost similar to countries that achieved fairly good containment (France, Spain, Germany, Italy), and is much better than UK, USA or Sweden. These results imply that the social distancing is the largest factor to achieve containment, and the contribution of broad PCR tests is smaller.

## Conflict of Interes

None reported.

## Data Availability

Prof. Masahiro Sonoo, the corresponding author, has access and availability of all the data referred in the manuscript.

## Acknowledgement

We would like to thank Dr. Katsuhisa Ogata, Department of Neurology, Higashisaitama National Hospital for the information on world countries.

